# Making Accelerating Medicines Partnership Data Findable and Interoperable through a Common Data Model: Extending OMOP for Multi-Source Multimodal Data

**DOI:** 10.64898/2026.08.31.26361831

**Authors:** Cole Tindall, Rodney Alan Long, Bart Naughton, Brandy M. Mapes, Dave Vismer, Halcyon G. Skinner, Jessica Malenfant, Mano R. Maurya, Mike A. Nalls, Srinivasan Ramachandran, Trang Nguyen, Mette A. Peters, Richard H. Scheuermann

**Affiliations:** DataTecnica LLC, Washington, DC; Eisai Inc, Cambridge, MA; Vanderbilt University Medical Center, Nashville, TN; Technome, Washington, DC; Verily Health, San Bruno, CA; Sage Bionetworks, Seattle, WA; University of California San Diego, La Jolla, CA; Broad Institute, Cambridge, MA; Division of Intramural Research, National Library of Medicine, National Institutes of Health, Bethesda, MD

**Author notes:** These authors contributed equally to this work (Cole Tindall, Rodney Alan Long).

## Abstract

SysBio FAIRplex is a Common Fund Venture Program^1^ that catalogs and indexes data from the Accelerating Medicines Partnership® (AMP®) Program^2^ through a federated model in which data hosts retain custody of their datasets. The central piece of this work is the SysBio Common Data Model (SysBio CDM). AMP is a precompetitive public-private partnership started in 2014 that unites the resources of NIH and private partners to improve our understanding of disease pathways and transform current models for developing new treatments by:

- identifying new targets, biomarkers, and development paradigms;
- developing leading-edge tools and technologies;
- collecting large-scale datasets and supporting analytics for open analysis by the public; and
- generating consensus platforms and procedures.

A multidisciplinary Task Force was chartered to design the SysBio CDM by extending the Observational Medical Outcomes Partnership (OMOP) Common Data Model^3^ into the -omics domain. The Task Force produced a Minimum Viable Product comprising nine OMOP tables; four extension tables for assay and file metadata; and a Common Data Element (CDE) Registry to specify field semantics. This manuscript describes the deliverable: the underlying design choices, the criteria applied in selecting and constructing the extension tables, how the extended model supports multimodal data integration across AMP projects, and what further work to support additional -omics modalities would entail. As an auxiliary methodology, the paper also describes the AI-assisted CDE harmonization workflow used to populate the model.

## Introduction

The Accelerating Medicines Partnership® (AMP®) program has, over more than a decade, generated high-value, disease-specific datasets across neurological, immunological, metabolic, and cardiovascular disease domains. These datasets are produced under pre-competitive principles intended to make data broadly accessible to the research community (Appendix A). In practice, however, AMP data are siloed across project-specific portals with differing schemas, vocabularies, and conventions. This fragmentation limits cross-disease query, integrative analysis, and systems-level discovery - particularly for -omics data, which now constitutes a growing share of AMP outputs.

SysBio FAIRplex (a Findable, Accessible, Interoperable, Reusable [FAIR] Platform for Exploration of Systems Biology) addresses this challenge by harmonizing bulk -omics and accompanying tabular metadata (clinical, observational, biospecimen, and assay descriptors) and by providing a federated platform for the discovery and use of AMP-hosted data (Figure 1). The cornerstone of this effort is the SysBio Common Data Model (SysBio CDM). To design the CDM, SysBio FAIRplex engaged a multidisciplinary Task Force. This manuscript is the Task Force’s report on its central deliverable: the SysBio CDM as an extension of an established CDM into the -omics domain.

**Figure 1.**
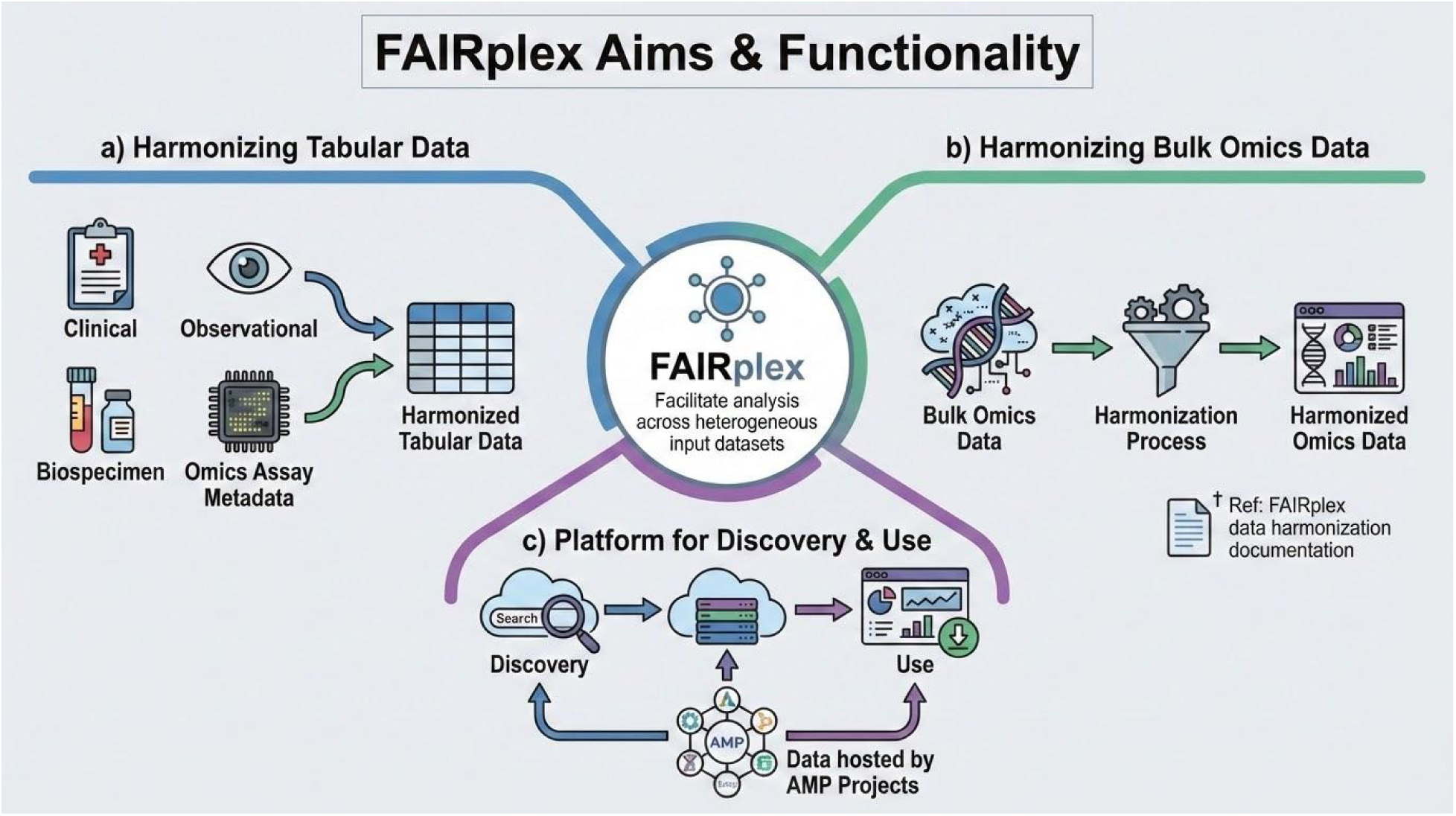
SysBio FAIRplex facilitates analysis across heterogeneous input datasets through three efforts: (a) harmonization of tabular data (clinical, observational, biospecimen, and -omics assay metadata) to a common data model; (b) harmonization of bulk -omics data; and (c) provision of a platform for the discovery and use of data hosted by AMP projects.

### Task Force Approach and Scope

The Task Force was charged with designing a CDM that allows tabular metadata from AMP projects to be represented in a single shared structure for cross-disease discovery and analysis, without requiring data hosts to move or reformat their data. It comprised Subject Matter Experts (SMEs) in AMP project data structures and data modeling, representatives from the NIH Common Fund, and AMP private partners.

The Task Force conducted a focused sprint to design a SysBio CDM Minimum Viable Product (MVP) grounded in 11 datasets from four AMP projects spanning neurological, autoimmune, and metabolic disease domains: AMP AD^4^, AMP CMD^5,6^, AMP PDRD^7^, and AMP RA/SLE^8^ (Appendix B). Although the SysBio CDM must ultimately scale across many assay types and disease areas, anchoring the MVP in a representative subset gave the Task Force a tangible target while exercising the model against the heterogeneity that any CDM adoption must accommodate:

- Multiple disease areas (Alzheimer’s disease, Parkinson’s disease, common metabolic diseases, autoimmune diseases).
- Antemortem versus postmortem assessments.
- Cross-sectional versus longitudinal study designs.
- A variety of biospecimen sources (brain, kidney, liver, adipose, muscle, hypothalamus, urine).
- Different schemas, formats, and data dictionaries across projects.
- Non-uniform vocabularies for diseases, phenotypes, tissues, and assays.
- Assay-specific terminology differences across portals.
- Different units of measurement and value sets for equivalent metadata elements.
- Mixed representation of metadata - some as structured tables, some as unstructured documents (PDFs and portal website text).

### Extending OMOP: Gap Analysis and Rationale

A key Task Force recommendation was to adopt an existing CDM as the framework rather than build one from scratch. The Task Force selected the OMOP CDM developed by the Observational Health Data Sciences and Informatics (OHDSI)^9^ community for several reasons: it is an extensive open-source relational standard for uniformly structuring observational healthcare data; OHDSI provides active user support, working groups, and a comprehensive open-source tool suite; and OMOP enjoys adoption by more than 4,700 investigators across 85+ countries, including major programs such as *All of Us*^10^ and the eMERGE Network^11^.

Despite OMOP’s strengths, the Task Force identified critical gaps in its native scope when applied to AMP -omics data:

- **Assay metadata.** OMOP was designed primarily for observational clinical data - encounters, diagnoses, medications, and procedures. It lacks structures to capture how an assay was performed, which is essential for interpreting and comparing -omics measurements across studies and sites.
- **File storage formats.** OMOP’s relational model is not designed for the array-based or file-based storage formats typical of -omics data (e.g., VCF, FASTQ, HDF5). Put plainly: OMOP can describe who the patient is and what their clinical history is, but cannot formally describe how -omics data were generated or where the resulting data files reside.
- **Multi-source semantics.** OMOP provides flexibility to use different semantics (e.g. units, ontologies) for individual fields. That works well when all data comes from one source, but relies on unspecified mechanisms to ensure new data, potentially from other sources, is consistent.

These gaps defined the extension problem the Task Force set out to solve. Rather than abandoning OMOP for an -omics-specific model - which would have sacrificed OMOP’s mature tooling, vocabularies, and community - the Task Force chose to extend OMOP with purpose-built tables that link to OMOP entities through standard relational mechanisms, and with a CDE Registry to make field semantics explicit.

### The SysBio CDM: An OMOP Extension for -Omics Data

The OMOP CDM (v5.4)^12^ organizes data into 36 relational tables grouped into six major data categories, each serving a distinct functional role within the model. The SysBio CDM MVP uses 9 of these OMOP tables and adds 4 custom extension tables purpose-built for -omics data, giving a total of 13 tables in the MVP. The Entity Relationship Diagram (ERD) in Figure 2 defines entities, primary and foreign key relationships, and cardinality constraints that together specify the conceptual framework for database design, data integration, and interoperability across heterogeneous data sources.

**Figure 2.**
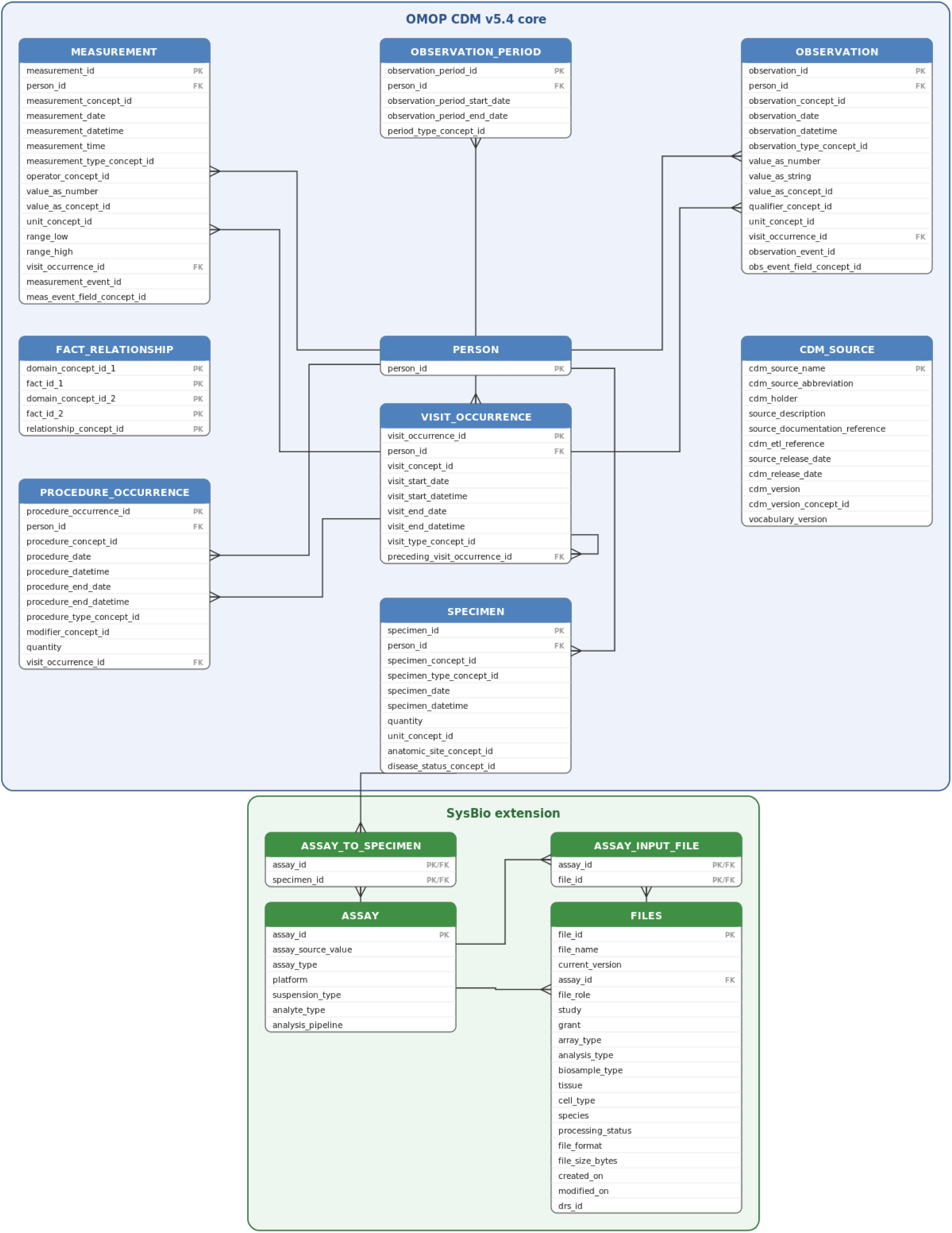
Detailed view of an ERD for the SysBio CDM. The core OMOP CDM v5.4 tables (blue) are extended by the SysBio ASSAY and FILE tables along with their corresponding lineage tables (green). Primary keys are labeled PK and foreign keys labeled FK. The crow’s foot marks indicate the ‘many’ side of a relationship. Foreign keys within OMOP CDM tables are excluded from this diagram if the joined table is unused in SysBio CDM. Source value fields are excluded from the diagram as they are fairly consistent across tables.

#### OMOP Tables Used in the SysBio CDM

The SysBio CDM uses the following 9 OMOP tables to represent participants, their clinical context, and their biospecimens:

- **PERSON** - Central record of individual study participants. Demographic information is stored in OBSERVATION instead of here, to allow for record-level governance.
- **OBSERVATION** - Clinical information output from observations or procedures. Used to document health-related events such as comorbidities and to further qualify disease status. Demographic data is stored here using existing OMOP demographic concepts/entities (e.g. Race (3046853), Ethnicity (40771985), Sex (3046965)).
- **PROCEDURE_OCCURRENCE** - Diagnostic or therapeutic procedures performed on a person. Used to record specimen collection procedures and broad anatomical site, linked to a study timepoint.
- **VISIT_OCCURRENCE** - Study visits. Used to anchor data to baseline and follow-up timepoints.
- **SPECIMEN** - Biological samples from a person. Used to record specimen IDs and granular anatomical site.
- **MEASUREMENT** - Numerical or categorical values obtained from examinations or sample testing. Used to record standardized assessment and instrument outputs. Specimen source concepts and measurement event fields (measurement_event_id) are used to tie a measurement to its source specimen, providing specimen lineage to a degree^13^.
- **OBSERVATION_PERIOD** - Denotes the time period in which the participant’s data was obtained. When defined calendar dates are unavailable, sentinel dates will be used to represent the time duration in which data was captured^14^.
- **FACT_RELATIONSHIP** - A generic associative entity table which applies standardized labels to describe the relationship between two records. The domain_concept_id fields define the tables and the fact_id fields specify the record. Often used to enrich or contextualize data by linking related records.
- **CDM_SOURCE** - Single provenance row to link to contributing AMP source; references the CDE Registry for the data-element definitions and ETL.

#### OMOP Extension Tables: ASSAY and FILES

The four extension tables address the identified OMOP gaps and are a principal technical contribution of the Task Force.

- **ASSAY** - Captures metadata about the -omics assays themselves: assay type, platform, protocol identifiers, and other attributes necessary to interpret and compare results across studies. ASSAY is linked to FILES (associating an assay execution with its outputs), to OMOP SPECIMEN (tying an assay to the biospecimen it was performed on), and to OMOP CDM_SOURCE (for provenance). This linkage pattern keeps assay metadata in a domain-specific table while preserving OMOP’s native handling of clinical and specimen context^15^.
- **FILES** - Captures information about data files and how to access them (file links, identifiers, formats, and metadata for federated retrieval). FILES is linked to ASSAY (associating files with the assay that produced them), to OMOP VISIT_OCCURRENCE (anchoring files to a visit context when applicable), and to OMOP CDM_SOURCE.
- **ASSAY_TO_SPECIMEN** - Connects the particular assay(s) used in -omics generation to the biosample.
- **ASSAY_INPUT_FILE** - Due to the per sample nature of many inputs in generating -omics data this link is necessary provenance for reprocessing.

#### Criteria the Task Force Applied in Selecting and Constructing the Extension Tables

The Task Force applied the following five criteria when deciding what to add as a custom table versus what to map into existing OMOP tables:

1. **Native fit first.** Any concept that could be faithfully represented in an existing OMOP table was mapped there. Custom tables were reserved for concepts OMOP cannot natively express.
2. **Minimize the extension surface.** The Task Force converged on the smallest extension that satisfied the immediate -omics requirements. Two tables plus two lineage tables, rather than a sprawling -omics schema, reduce maintenance burden and preserve compatibility with OMOP tooling.
3. **Link, do not modify.** Extension tables connect to OMOP through foreign keys to existing OMOP entities. No OMOP table definitions are altered, so OMOP-compatible tools and vocabularies continue to apply.
4. **File metadata, not file payloads.** FILES stores pointers and descriptors, consistent with the federated FAIRplex model in which data physically resides with AMP hosts.
5. **Extensibility.** The ASSAY table was designed so that additional assay types and platform-specific attributes can be added without restructuring the rest of the model.

#### How These Choices Support Multimodal Data Integration

By keeping participant- and specimen-level data in OMOP and confining -omics-specific structure to ASSAY and FILES, the SysBio CDM enables cross-modal queries that join clinical state with assay-level evidence using standard relational operations. A query such as "*find all single-cell RNA-seq files from kidney specimens of participants diagnosed with lupus*" traverses CONDITION_OCCURRENCE → PERSON → SPECIMEN → ASSAY → FILES along well-defined keys. The extension thus turns multimodal integration from a complex integration project into a query. (Note that reducing the integration burden will not fully eliminate the need to assess scientific comparability and fitness-for-use via HITL review by SMEs.)

#### The SysBio CDM MVP Schema and Reference Implementation

The complete schema underlying the CDM itself, including field-level definitions and value sets, is maintained in machine-readable form in the FAIRplex CDM GitHub repository as a reference resource^16^. The repository serves as the canonical source for the CDM and as the implementation framework to operationalize the CDM when the CDM is deployed in the FAIRplex production environment. A changelog tracks all schema changes and additions as the CDM evolves.

### Applying the Extended CDM Across AMP Disease Domains

To demonstrate that the extension would support cross-domain integration, the Task Force exercised the MVP against 11 datasets from four AMP projects (Appendix B). The datasets span four disease areas, multiple tissues (brain, liver, adipose, muscle, hypothalamus, kidney, urine), and both cross-sectional and longitudinal designs. Each dataset was modeled through the same OMOP backbone, with assay metadata and file pointers represented in the ASSAY and FILES extension tables. The resulting integrated representation enables queries that span disease, tissue, and assay type with no project-specific schema knowledge required of the analyst.

#### Worked Example: Looking for Shared Neurodegeneration Signatures Across AMP AD and AMP PDRD

A neuroscientist wants to test whether genes that are upregulated in postmortem brain tissue from Alzheimer’s disease (AD) participants show similar expression changes in Parkinson’s disease (PD) brain tissue, as a way of asking whether the two diseases share molecular features. Without a harmonized CDM, this analysis would require the investigator to learn the AMP AD and AMP PDRD portal schemas, manually reconcile participant- and specimen-level descriptors, normalize assay metadata that uses different field names in each portal, and download files individually from each host - a multi-week exercise before any analysis begins.

With the SysBio CDM, the same question becomes a single query expressed against a shared structure. The investigator filters PERSONs by CONDITION_OCCURRENCE for Alzheimer’s disease or Parkinson’s disease (using the OMOP CONCEPT_ID for each, drawn from SNOMED CT). They retrieve the associated SPECIMENs and restrict to brain tissue via the granular anatomical-site field. They follow each SPECIMEN to its ASSAY records and restrict to bulk RNA-seq. Finally, they follow the ASSAY records to FILES to obtain file pointers to the originating AMP host.

The CDE Registry guarantees that "Alzheimer’s disease" means the same thing in both projects, that brain anatomical sites are mapped to a common vocabulary, and that the assay descriptors are comparable. The integration has become a query against a shared structure rather than a per-project translation problem - exactly the cross-disease analysis the AMP program was designed to enable but that data siloing has historically frustrated.

### Extending the CDM Further: Additional -Omics Modalities and Future Work

The MVP was scoped to single-cell RNA-seq (scRNA-seq) because these assays were available across all four contributing AMP projects. The same extension pattern - ASSAY plus FILES, linked to OMOP through SPECIMEN, VISIT_OCCURRENCE, and CDM_SOURCE - is intended to accommodate additional -omics modalities with modest, well-bounded changes. The Task Force identified the following directions for further extension:

- **Additional bulk and single-cell assays.** Proteomics, metabolomics, lipidomics, epigenomics, and bulk and spatial transcriptomics can be added by extending the assay-type vocabulary used in the ASSAY table and adding any assay-class-specific fields necessary to interpret the data. The relational structure does not need to change.
- **Spatial transcriptomics and sample derivatives.** AMP AIM, for example, anticipates releasing spatial transcriptomics datasets in which multiple RNA samples are isolated from a single tissue section. Capturing such derivative relationships is an open design question: it can be addressed within the OMOP SPECIMEN table using existing concepts, by developing new OMOP concepts for biospecimen derivatives, or by introducing a lightweight specimen-derivative extension^17^. The Task Force recommends evaluating these options in the next sprint.
- **Subcellular location, pseudotime, and other emerging dimensions.** As experimental technologies evolve, additional axes (e.g., subcellular compartment, pseudotime ordering) can be incorporated as CDEs and represented through the existing extension tables. Related collaborative efforts, such as the Path-NeuroDegeneration consortium^18^, are building provisions for this growing and scalable integration of -omics and imaging modalities; that consortium will release over 1,000 updated CDEs for brain neuropathology and spatial transcriptomics focused on neurodegenerative diseases.
- **Imaging, sensor, and high-dimensional longitudinal data.** OMOP is well suited to harmonizing clinical records, diagnoses, medications, procedures, and some derived molecular or laboratory measurements, but raw, high-dimensional, longitudinal -omics, imaging, and sensor datasets are beyond its native scope. A more extensive hybrid strategy - potentially involving additional extension tables analogous to ASSAY/FILES for imaging or sensor data - may be required.
- **Vocabulary expansion.** Each OMOP record references a CONCEPT_ID linked to controlled vocabularies (SNOMED CT, RxNorm, LOINC, and others). The Task Force identified additional vocabularies and ontologies of relevance to SysBio that will be evaluated for inclusion in future sprints.

In each case, the question is not whether the SysBio CDM can be extended further - the extension pattern is deliberately designed to support exactly this - but rather which specific extensions deliver the most cross-domain analytical value for AMP and the broader community.

### Auxiliary Methodology: AI-Assisted CDE-Based Harmonization

The preceding sections describe the SysBio CDM itself. This section describes the methodology the Task Force chose for populating it. The chosen approach combines the CDM and associated CDE Registry with an AI-assisted Human-In-The-Loop (HITL) harmonization workflow. The CDE registry serves as the substrate for the harmonization phase, a Rosetta Stone for data elements across resources that has been centrally parameterized to help govern content. The methodology is described here in summary; full implementation details are provided in the Supplementary Material.

#### The CDE Registry Layer

While OMOP provides a robust storage model, it does not by design enforce consistent value sets, units, or encodings across independently harmonized datasets. The CDE Registry sits between source AMP variables and OMOP storage, ensuring that variables representing equivalent concepts map to identical target representations and reducing the semantic drift that arises from independent ETL (Extract-Transform-Load) projects. Table 1 contrasts OMOP’s role as a storage format with the CDE’s role as specification. CDEs from this project can be found in a natural language searchable database within the SysBio FAIRplex portal.

**Table 1.** Storage Format versus Specification: OMOP and CDE Attribute Comparison.

| Attribute | OMOP (Storage) | CDE (Specification) |
| --- | --- | --- |
| Value set | Accepts any valid concept | Defines concept value set |
| Units | Accepts any valid unit | Defines a common unit for harmonized data |
| Encoding | Accepts numeric OR string | Defines which to use |
| Non-OMOP questions or surveys | Not stored | Preserved |
| Multi-concept patterns | Stores individual concepts | Defines how concepts combine |

The CDE Registry allows researchers to do fitness-for-use assessment (CDE existence, AMP coverage, and per-AMP qualifier availability) before requesting data. It does this by incorporating value set normalization, capture of context that lives in protocols rather than data dictionaries, and derivative flagging (granular variables derive general queries). These functions are illustrated in Figure 3.

**Figure 3.**
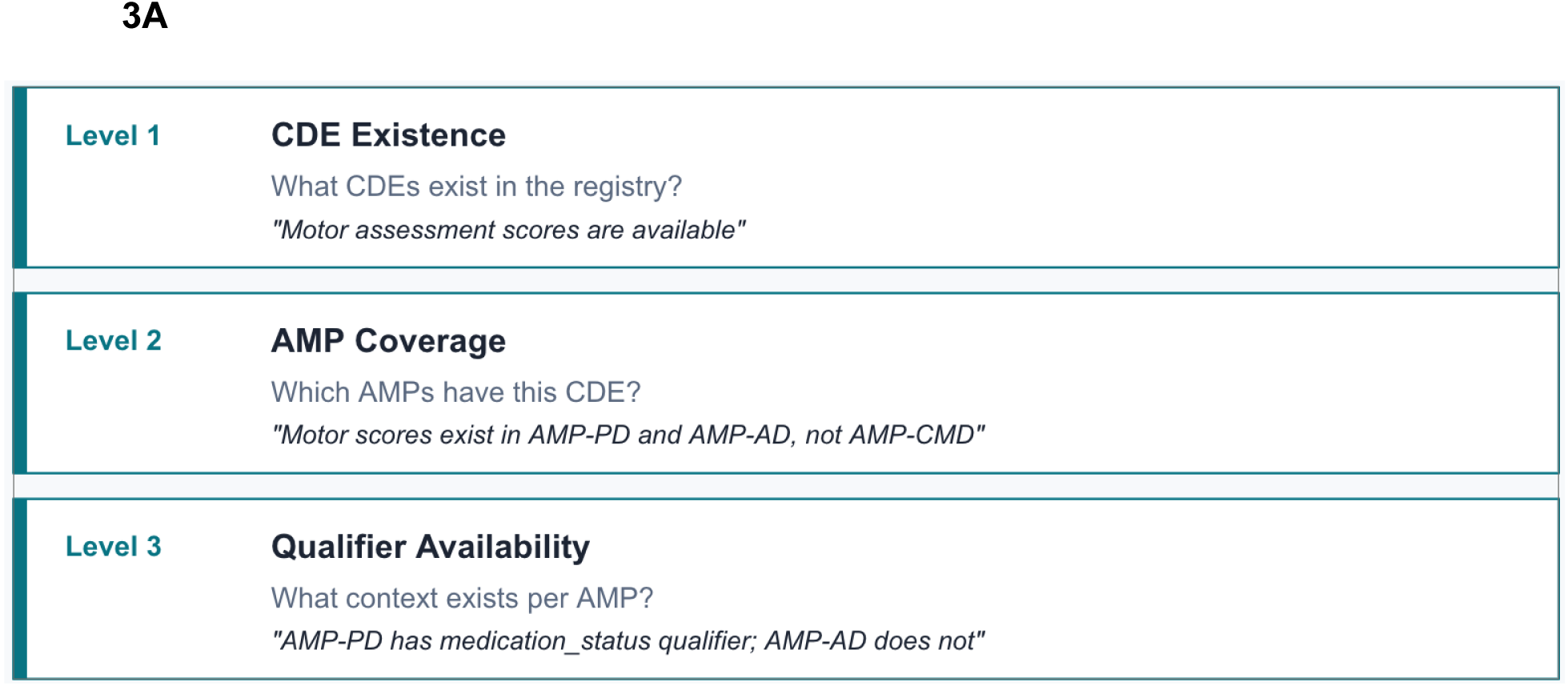
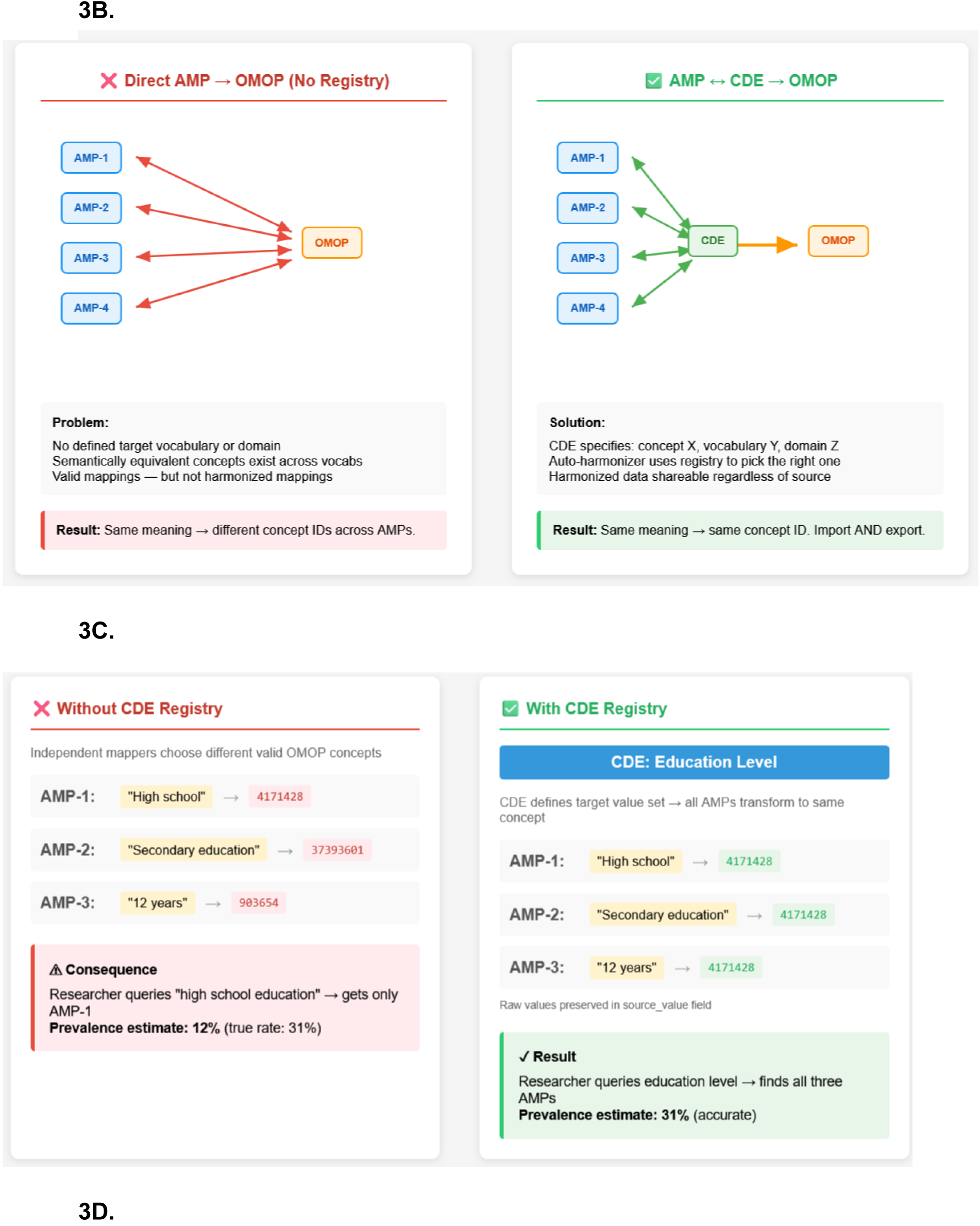
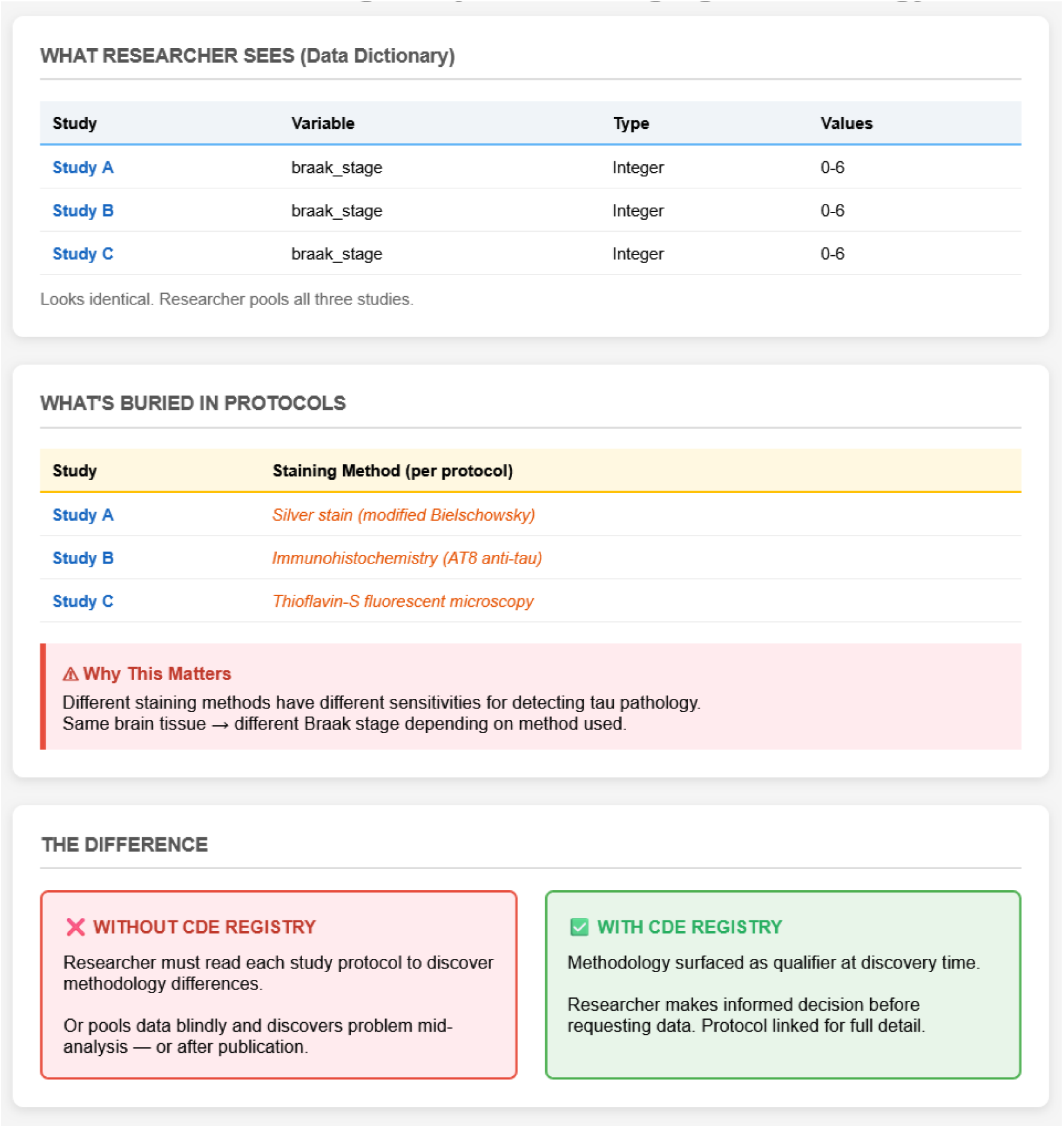
CDE Registry architecture and value. (A) Three levels of informed data selection: CDE existence, AMP coverage, and qualifier availability. (B) Single point of change: AMP→CDE→OMOP transformation versus direct AMP→OMOP mapping. (C) Value set normalization: the CDE defines the target concept ID across all sources. (D) Hidden heterogeneity: Braak staging appears identical across studies (integer 0–6) but reflects different staining methods with different sensitivities for tau pathology detection.

#### The AI-Assisted HITL Harmonization Workflow

For each AMP data element, the FAIRplex CDE harmonization process^19^ maps the element to a CDE library (prioritizing the NIH CDE repository^20^), translates data values to be consistent with the mapped CDE, and assigns the element to the appropriate OMOP table. The harmonization addresses fundamental limitations of direct automated mapping to representations such as OMOP, ICD-10, and other structured vocabularies, which can be time-consuming, error-prone, tuned for storage rather than specification, and inflexible for -omics or disease-specific metadata. A HITL review by domain experts on the Task Force adjudicates AI recommendations - accepting, rejecting, or modifying mappings - before they are committed to the CDE Registry and used to populate the OMOP tables. A representative set of clinical variables was reviewed by the Task Force as a whole; the full set of mappings from the 11 input datasets was reviewed by a smaller Task Force administrative team, applying Task Force recommendations where applicable. Versioning via DOI and updates to the CDEs will be tracked within the FAIRplex portal and quarterly updates to the open source reference corpus on Hugging Face^21^ as a SysBio CDM collection (the entire corpus can be downloaded at^22^, with a special collection tagged SysBio with AMP as the source).

Each harmonization run produces a harmonization record (Appendix C) that tracks input source variables, source files, and how they were mapped, supporting provenance and source-AMP verification.

#### Why This Methodology Was Chosen

Direct, fully automated source-to-OMOP mapping is fast but error-prone and inflexible for -omics-specific and disease-specific metadata. Fully manual mapping is accurate but does not scale to AMP’s volume. The CDE Registry plus AI-assisted HITL approach combines automated draft mappings with expert oversight at decision points, producing scalable, auditable harmonization that preserves semantic fidelity.

#### Future Direction of the CDE Registry: Parameterized CDEs, JSON Schema, and NIH Integration

To support interoperability across AMP, the Common Fund Data Ecosystem (CFDE), and other data resources, SysBio FAIRplex plans to develop parameterized CDEs for clinical and sample metadata associated with -omics data. Each CDE will be cataloged so that variables representing equivalent information content share identical headers and conform to standardized numeric or categorical scales. This harmonization applies to existing repositories and establishes a specification for related datasets that may be integrated in the future, thereby reducing barriers to cross-cohort analysis and enhancing AI readiness through consistent feature representation.

All CDEs will be transformed from flat tabular formats to JSON schema for integration with the NIH CDE Repository. Overlap with existing NIH-validated CDEs and OMOP concept terms will be noted. This transition from human-readable specifications to machine-readable, validatable schemas supports the FAIR principles by improving findability and reusability while lessening the time analysts must spend on data formatting when working across federated datasets. The NIH CDE Repository will serve as the registry of note for this growing, evolving collection of validated CDEs, facilitating central browsability and attribution to the AMP stakeholders.

## Discussion

This work delivers an OMOP extension - the SysBio CDM - that brings -omics assay metadata and file linkages into a CDM framework that already enjoys broad clinical-data adoption. The extension is deliberately small (two tables plus two lineage tables) and link-based rather than invasive, which preserves OMOP tooling and lowers the barrier for any institution familiar with OMOP to adopt the model. Within the AMP Program, where disease-specific portals have historically evolved independently, the SysBio CDM provides a unifying semantic layer that enables cross-project querying and integrative analysis across diseases, tissues, and molecular modalities.

The extension’s value rests on three Task Force design choices: anchoring -omics data to OMOP entities through standard relational links, keeping the extension surface small, and supporting the model with a CDE-based specification layer that enforces consistent semantics during ETL. Together, these allow multimodal integration to be expressed as a query rather than as a bespoke integration project.

The SysBio CDM also has utility for prospective data harmonization. Rather than treating harmonization purely as a downstream ETL problem, the CDM and CDE Registry position harmonization as a forward-looking specification for how information should be represented, encoded, and qualified across studies. As new AMP projects come online, prospective adoption of SysBio CDEs can reduce future harmonization effort by ensuring newly generated data is interoperable from the outset. Parameterized CDEs provide flexibility to standardize core constructs (demographics, disease staging, biospecimen attributes, assay descriptors) while still allowing domain-specific extension - balancing cross-project comparability against the reality that different disease domains and assay platforms require tailored metadata.

### Limitations

The SysBio CDM was exercised across multiple AMP disease domains, but the MVP implementation reflects a limited subset of available datasets and -omics platforms. Extension to additional assay types, emerging modalities, and more heterogeneous study designs will require ongoing schema refinement and validation, and the CDE Registry will need to evolve as new technologies emerge. The AI-assisted mapping workflow accelerates harmonization but does not eliminate the need for expert oversight; the observed rate of expert edits underscores that automated semantic matching remains imperfect for nuanced clinical constructs, assay-specific variables, sparse input data dictionaries, and context-dependent measurements.

### Open Items and Next Steps

The Task Force identified the following items as open and recommended for future work: (1) testing the SysBio CDM throughout the FAIRplex ecosystem - from search tools through data retrieval - including access-control checks so that only variables a user is approved to access are returned by queries; (2) deciding the representation of sample derivatives, particularly for spatial transcriptomics where multiple samples may derive from one tissue section; (3) evaluating the additional controlled vocabularies and ontologies surfaced by the Task Force for inclusion in the OMOP CONCEPT_ID space; (4) extending the model and CDE Registry to assay types beyond RNA-seq/scRNA-seq; and (5) versioned release notes and provenance tracking within the SysBio portal and connected documentation at implementation of the model.

## Conclusion

The SysBio CDM MVP demonstrates that the OMOP Common Data Model can be extended into the -omics domain through a small, link-based extension without modifying OMOP itself. Combined with a CDE specification layer and an AI-assisted HITL harmonization workflow, the resulting framework enables consistent semantics, interoperable metadata, and fitness-for-use assessment across federated AMP datasets. The resulting CDM and CDE Registry provide durable infrastructure aligned with the FAIR principles, lowering barriers to cross-disease discovery, systems-level analysis, and AI-ready data integration. The extension pattern established in this MVP is intended to support further -omics modalities as the model matures. While interoperability and discovery are enabled through the CDM, governance, access controls, and source-system policies remain enforced by participating AMP resources. As AMP and related initiatives continue to expand in scale and modality, prospective adoption of shared data specifications via the SysBio CDM offers a path to reducing harmonization effort and maximizing the long-term scientific value of large, multi-institutional biomedical data assets.

## Data Availability

Code and resources generated in this study are contained in the manuscript
All data used in this manuscript are available online at:
- AD Knowledge Portal /AMP AD Program
- AMP CMD/CMDGenome Atlas
- AMP PDRD
- ARK Poral/AMP RA/SLE Program

https://huggingface.co/datasets/DataTecnica/RoP_biomedical/tree/main/v2026.08

https://github.com/SysBio-FAIRplex/SysBio-CDM

https://adknowledgeportal.synapse.org/Explore/Programs/DetailsPage?Program=AMP-AD

https://cmdga.org/

https://amp-pdrd.org

https://arkportal.synapse.org/Explore/Programs/DetailsPage?

## Acknowledgements

This work was supported by the SysBio FAIRplex Initiative, a FAIR PLatform for EXploration of Systems Biology, funded by the National Institutes of Health, Office of the Director: Common Fund Other Transactions Programs: 1OT2OD037975. In addition, SysBio FAIRplex would not be possible without the partnership of the Accelerating Medicines Partnership® (AMP®) program and the Foundation for the National Institutes of Health (FNIH).

We thank Brandy A. Quinn, Technome, Washington, DC, for administrative support for the SysBio CDM Task Force.

## Appendix

## Appendix A: Active and Completed AMP Projects

| AMP Project | Status | Disease Area | Primary Focus | Major Data Types |
| --- | --- | --- | --- | --- |
| AMP AD (1 & 2) | AD1: Completed;<br>AD2: Active | Alzheimer's Disease | Target and biomarker discovery via deep multi-omic and clinical data | Raw and processed clinical, genetics, transcriptomics (bulk and single-cell), epigenomics, proteomics, metabolomics/lipidomics. |
| AMP AIM | Active | Autoimmune & Immune-Mediated Diseases | Deep cellular and spatial mapping of autoimmune mechanisms | Clinical, genetics, and spatial transcriptomics (planned). |
| AMP ALS | Active | Amyotrophic Lateral Sclerosis | Biomarker and target discovery; progression modeling | Individual-level epigenomics, transcriptomics, and clinical data including longitudinal cohorts. |
| AMP BGTC | Active | Bespoke Gene Therapy Consortium (rare genetic diseases) | Platforms and standards for tailored gene therapies | Data from gene therapy development, evaluation, and trials. |
| AMP CMD | Active | Common Metabolic Diseases | Genetic and functional drivers of metabolic diseases | Aggregate summary data (genetics, transcriptomics, epigenomics); individual-level processed transcriptomics and epigenomics. |
| AMP HF | Active | Heart Failure | Phenotyping and precision medicine framework | Clinical, genetics, transcriptomics, epigenomics, proteomics, metabolomics, imaging (planned). |
| AMP PDRD | Active | Parkinson's Disease and Related Disorders | Biomarker discovery and subgroup characterization | Clinical, genetics, transcriptomics, proteomics from disease target and peripheral tissue. |
| AMP RA/SLE | Completed | Rheumatoid Arthritis and Systemic Lupus Erythematosus | Single-cell and multi-omic disease deconstruction (foundation for AMP AIM) | Clinical, genetics, transcriptomics, epigenomics, proteomics, cytometry. |
| AMP SCZ | Active | Schizophrenia | Mechanism and early-risk characterization | Clinical, behavioral, neurophysiology, neuroimaging, wearables, interview transcripts. |
| AMP T2D | Completed | Type 2 Diabetes | Therapy-development resources (foundation for AMP CMD) | Aggregate summary data and individual-level processed transcriptomics and epigenomics. |

## Appendix B: AMP Projects and Datasets Included in the SysBio CDM MVP

| AMP Project | Dataset Name | Tissue | Study Type |
| --- | --- | --- | --- |
| AMP AD | AMP-AD_DiverseCohorts | Brain | Cross-sectional |
| AMP AD | ROSMAP | Brain | Cross-sectional |
| AMP AD | MIT_ROSMAP_Multiomics | Brain | Cross-sectional |
| AMP CMD | High-resolution multi-omic analysis of liver cells | Liver | Cross-sectional |
| AMP CMD | Single-cell atlas of human visceral and subcutaneous adipose tissue | Adipose | Cross-sectional |
| AMP CMD | Genetic regulation in human skeletal muscle | Muscle | Cross-sectional |
| AMP CMD | Spatio-cellular map of the human hypothalamus | Hypothalamus | Cross-sectional |
| AMP CMD | Atlas of healthy and injured cell states and niches in the human kidney | Kidney | Cross-sectional |
| AMP PDRD | Post-mortem sequencing cohort | Brain | Cross-sectional |
| AMP RA/SLE | AMP SLE Phase II Urine scRNA-seq | Urine | Longitudinal |
| AMP RA/SLE | AMP SLE scRNASeq | Kidney | Cross-sectional |

## Appendix C: FAIRplex Harmonization Record Description

| Column Name | Description |
| --- | --- |
| Source(s) | AMP Project source of input variables (AMP-AD, AMP-CMD, AMP-PD, AMP-RA/SLE). |
| Source_File | File for tracking provenance from where the variable was ingested. |
| Source_AMP_Variable | Original source field name as it appears in the raw AMP dataset prior to harmonization. |
| AMP_Variable_Description | Description of the source variable as documented in the AMP metadata file and source data dictionary. |
| match_method | Method used by the AI harmonizer to suggest CDE matches for the input variable. |
| match_status | Outcome of the matching process: high_quality, medium_quality, or needs a new CDE. |
| matched_description | Description of the meaning of the match as interpreted during review (contextualized semantic explanation). |
| matched_collection | CDE library, ontology, or domain to which the match belongs (e.g., NINDS CDE, AMP-RA CDE library, internal catalog). |
| rationale | Justification describing why the match was selected, rejected, or flagged. |
| alternative_matches | Additional candidate CDEs identified during the search process. |
| needs_new_cde | Boolean (TRUE/FALSE). Indicates that no existing CDE adequately represents the variable. |
| recommended_action | Next step for resolution (e.g., accept_match, generate_new_cde). |
| Matched_CDE | Name of the selected existing CDE, if a match was found. |
| Matched_CDE_Collection | Collection or registry that defines the matched CDE. |
| generated_CDE | New CDE generated if a match was not found. |
| TinyID | URL for the NLM CDE repository entry for the matched CDE. |
| OMOP_Concept_Name | Standard OMOP clinical concept name associated with the mapping, when applicable. |
| OMOP_Concept_ID | OMOP numeric concept identifier supporting standardized analytics and interoperability. |

## Appendix D: Glossary of Terms and Acronyms

| Term / Acronym | Definition |
| --- | --- |
| AMP | Accelerating Medicines Partnership; a public–private partnership funding disease-specific consortia. |
| CDM | Common Data Model; a shared structural specification for data so that independently produced datasets can be combined. |
| CDE | Common Data Element; a standardized definition of a single data field, including allowed values, units, and encoding. |
| CONCEPT_ID | An OMOP identifier that links a stored value to a standardized concept in a controlled vocabulary. |
| ERD | Entity Relationship Diagram; a visual representation of database tables and the relationships between them. |
| ETL | Extract-Transform-Load; the process of pulling data from source systems, reformatting it, and loading it into a target schema. |
| FAIR | Findable, Accessible, Interoperable, Reusable; a set of principles for research data stewardship. |
| HITL | Human-In-The-Loop; a workflow in which human experts review and adjudicate machine-generated outputs. |
| MVP | Minimum Viable Product; a working version with enough functionality to validate the approach and inform further development. |
| OHDSI | Observational Health Data Sciences and Informatics; the open-science community that maintains OMOP. |
| -Omics | A class of high-throughput biological measurements covering many molecules at once (genomics, transcriptomics, proteomics, metabolomics, and others). |
| OMOP | Observational Medical Outcomes Partnership Common Data Model; the standard relational data model the SysBio CDM extends. |
| Ontology | A formal, structured vocabulary of terms and their relationships, used to make data computer-interpretable. |
| scRNA-seq | Single-cell RNA sequencing; an assay measuring gene expression in individual cells. |
| SME | Subject Matter Expert. |
| SNOMED CT, RxNorm, LOINC | Standardized clinical vocabularies referenced by OMOP CONCEPT_IDs (for conditions, drugs, and laboratory tests, respectively). |

